# Limits of Sociodemographic Data for Predicting Postpartum Readmission: Conventional, Ensemble and Automated Machine Learning Compared at Equal Workload, With a Decision-Economic Analysis

**DOI:** 10.64898/2025.12.16.25342440

**Authors:** Martin G. Frasch, Colin Wakefield, Ciprian P. Gheorghe

**Author notes:** **Corresponding Author:** Martin G. Frasch, MD PhD, Department of Obstetrics and Gynecology, Institute on Human Development and Disability, University of Washington School of Medicine, 1959 NE Pacific Street, Box 356460, Seattle, WA 98195-6460.

## Abstract

**Objective:** Postpartum readmission is rare and costly, and risk tools often rely on routinely collected sociodemographic data. We asked what such data can support, whether algorithm choice matters, and when a modest model could pay for itself.

**Methods:** We analyzed 8,774 participants of the nuMoM2b cohort of first pregnancies, of whom 154 (1.8%) were readmitted within 14 days. Conventional, ensemble and automated machine learning models (FLAML, AutoGluon) were trained on 70% of participants, with every fitted step confined to training data, and evaluated once on the remaining 30% (46 readmissions). We compared models at default thresholds and equal screening workload, after recalibration, across richer feature sets, and in economic scenarios against contacting all or no patients.

**Results:** The best model, a tuned logistic regression, reached an area under the ROC curve (AUC) of 0.614 (95% confidence interval 0.542–0.685). Both automated frameworks discriminated worse (AUC 0.416–0.519) after optimistic internal validation. Sensitivity at the default threshold ranged from 0% to 41% and reflected class-imbalance corrections rather than ranking ability; when every model flagged the same 28.4% of patients, conventional and ensemble models reached 11 to 20 of 46 readmissions. After recalibration, no model improved on predicting the base rate. Delivery and discharge information raised the AUC to 0.611–0.631, and learning curves were flat. Reweighted post hoc to each US state’s first births, the AUC ranged from 0.531 to 0.667, lowest in the least diverse states. Contacting everyone was optimal for low-cost outreach; near the break-even cost, no model gained more than its running cost.

**Conclusions:** Sociodemographic data alone cannot support a useful postpartum readmission tool, whatever the algorithm. Under class imbalance, default-threshold comparisons mislead. A modest model’s value depends on the population and the intervention, which decision-economic analysis, not discrimination alone, should assess. Future work should prioritize richer peripartum and postpartum data.

**Précis:** Sociodemographic data supported only modest prediction of postpartum readmission with any algorithm; apparent differences at the default threshold reflected probability scaling, and an economic analysis located the narrow range of intervention costs within which even a weakly discriminating model can add value.

**Highlights:**

1. Fourteen conventional, ensemble and automated machine learning models predicted 14-day postpartum readmission from sociodemographic data with modest discrimination (best test ROC-AUC 0.614); after recalibration none improved on predicting the base rate.
2. At the default 0.5 threshold, sensitivity ranged from 0% to 41% and tracked class-imbalance corrections; at an equal screening workload, conventional and ensemble models were indistinguishable.
3. Two AutoML frameworks discriminated worse on the test set than a regularized logistic regression because their internal validation was optimistic with 108 training events.
4. Delivery and discharge information raised ROC-AUC only to 0.611–0.631 and the learning curve was flat, pointing to richer peripartum and postpartum data rather than larger samples.
5. A model can add economic value only near the cost at which contacting all and none break even; with sourced cost and effectiveness inputs, no model’s gain there exceeded its running cost.
6. Transported post hoc to each US state’s first births, discrimination ranged from ROC-AUC 0.531 to 0.667 and was lowest in the least diverse states: a sociodemographic model’s value depends on the population it is applied to.

## 1 Introduction

Postpartum readmission represents a significant burden for new mothers, families, and healthcare systems, affecting approximately 1–2% of deliveries [1] and generating substantial inpatient costs. [2] Early identification of high-risk patients could enable targeted interventions to prevent readmissions, improve maternal outcomes, and reduce healthcare costs. However, developing effective prediction models requires a careful balance between model performance, clinical interpretability, and implementation feasibility.

The emergence of automated machine learning (AutoML) has promised to democratize predictive modeling in healthcare by automating complex processes of algorithm selection, hyperparameter optimization, and feature engineering. [3, 4] Leading AutoML frameworks such as FLAML and TPOT claim to deliver superior performance while reducing the need for specialized machine learning expertise. This technological promise has generated significant interest in healthcare applications, where AutoML could accelerate the development and deployment of clinical prediction tools.

However, the clinical utility of AutoML in healthcare settings remains largely unproven, and systematic reviews note that healthcare-specific challenges—particularly class imbalance and the need for calibrated, interpretable outputs—are not yet well handled by general-purpose AutoML systems. [5] Healthcare prediction models face unique challenges, including severe class imbalances, regulatory requirements for interpretability, computational resource constraints, and the critical importance of sensitivity over overall accuracy. [6, 7] Traditional machine learning approaches, particularly logistic regression, have demonstrated robust performance in clinical applications due to their interpretability, computational efficiency, and well-understood statistical properties. [8, 9]

The nuMoM2b (Nulliparous Pregnancy Outcomes Study: Monitoring Mothers-to-be) cohort is one of the largest prospective cohorts of first pregnancies. [10] Routinely collected sociodemographic information is available for almost every participant, as it is in most delivery units, so it is the natural starting point for a readmission risk tool. Whether it is sufficient is a separate question. A rare outcome also exposes an under-appreciated problem in comparing algorithms: under severe class imbalance, a model’s apparent sensitivity at the default 0.5 threshold depends on how resampling and class weighting shift its predicted probabilities, as much as on its ability to rank patients. [11]

We therefore set out to establish what routinely collected sociodemographic data can and cannot support for predicting 14-day postpartum readmission. Specifically, we asked (i) whether conventional, ensemble and automated machine learning (AutoML) differ in discrimination, calibration or clinical utility when compared at a common operating point rather than at their default thresholds; (ii) whether adding enrollment-interview or delivery and discharge information improves prediction; and (iii) at what intervention cost, if any, acting on a modest model’s predictions outperforms contacting every patient or no patient. Because negative and near-null results are informative for future data collection, we report every model variant we evaluated.

## 2 Methods

### 2.1 Study Design, Data Source and Outcome

This is a secondary analysis of the nuMoM2b prospective cohort of nulliparous women with singleton pregnancies, recruited at eight US clinical sites. [10] The outcome was rehospitalization within 14 days of delivery (chart-abstraction item CMAJ01), and participants with this item recorded formed the analyzed cohort. The unit of analysis was the participant; each participant contributed one delivery and one row. Reporting follows TRIPOD guidance. [9]

### 2.2 Candidate Predictors and Leakage Control

We classified all 11,617 codebook variables by their timing relative to the outcome (Supplementary Table S1). Primary analyses used 18 sociodemographic and pre-pregnancy candidates: race and ethnicity (four codings), body mass index and its category, education, gravidity, three smoking items, five insurance-payer items, income relative to the federal poverty level and its category. Two preprocessing rules governed how items were coded. First, items that are blank by design (cigarettes per day for participants who did not smoke in the three months before pregnancy; recent smoking for participants who had never used tobacco) were recoded as “none” rather than treated as missing. Second, unordered categorical items (race and ethnicity codings, marital status) were one-hot encoded, pooling categories with fewer than 20 participants, rather than entered as numeric codes.

Data were split once into training (70%) and test (30%) sets, stratified by outcome (seed 42); the code asserts that no participant appears in both. Every fitted step was learned on the training set or inside training folds and applied unchanged to the test set: imputation (median for numeric and ordinal items, mode for categorical items), standardization, recursive feature elimination (RFE; logistic-regression estimator, 10 features), SMOTE over-sampling, [12] hyperparameter search, decision thresholds and recalibration. The test set was used once per model, for final evaluation.

### 2.3 Models

#### Original configuration

The configurations specified at the start of the study, with the preprocessing rules above, were logistic regression (L2, *C* = 1, balanced class weights), random forest (100 trees, balanced class weights) and gradient boosting (scikit-learn defaults), each with RFE and SMOTE inside the pipeline; logistic regression was also fitted without SMOTE.

#### Tuned models

Logistic regression, random forest and gradient boosting were tuned by randomized search (40 draws, stratified 5-fold cross-validation (CV) within the training set, ROC-AUC). The search treated the imbalance strategy as a hyperparameter: SMOTE or none, and balanced or no class weights (Supplementary Table S4).

#### Ensembles

A stacking ensemble used the three base pipelines, with base-model probabilities from stratified 5-fold CV as inputs to a logistic-regression meta-learner (*C* = 1, balanced class weights). A soft-voting ensemble averaged the base models’ probabilities after isotonic calibration, each by CalibratedClassifierCV with stratified 5-fold CV within the training set. Each ensemble was built from both the original-configuration and the tuned base models.

#### Automated machine learning

FLAML [3] was run as originally configured (SMOTE-resampled training data, ROC-AUC, 300 s, default search space) and in an imbalance-aware configuration (original training data, balanced sample weights, 5-fold CV, ROC-AUC, 900 s). AutoGluon-Tabular [13] was run with the best quality preset (bagging and multi-layer stacking, ROC-AUC, 900 s). Both frameworks received the same fold-fitted imputation, encoding and scaling as the other models. Because both searches are bounded by wall-clock time, their results can differ between runs; we report the first complete run and, in Results, a full rerun of the pipeline.

### 2.4 Operating Points

For every model, out-of-fold (OOF) predictions were obtained for the training set by stratified 5-fold CV of the final configuration. For AutoGluon, whose internal OOF estimates are inflated by selecting among many candidate models on the same folds, OOF predictions came from refitting AutoGluon within each outer training fold (300 s per fold). Decision thresholds were selected on these OOF predictions only and then applied to the test set. They were the highest threshold reaching 80% and 70% OOF sensitivity, and the threshold flagging the same share of patients that the original-configuration logistic regression flagged at 0.5. This matched flag rate puts every model at an identical screening workload, so that their sensitivities can be compared directly. We also report each model at the default 0.5 threshold.

### 2.5 Calibration, Recalibration and Decision Curves

Calibration was summarized by the Brier score, the Brier score of a constant prediction equal to the training prevalence, and the calibration slope and intercept. For every model, Platt scaling (logistic regression on the logit of the OOF prediction) and isotonic regression were fitted to the training OOF predictions and applied to the test predictions. Decision curve analysis [14] compared the net benefit of recalibrated models with contacting all or no patients across threshold probabilities of 0.5–10%.

### 2.6 Statistical Analysis

Performance on the test set was summarized by ROC-AUC, PR-AUC (average precision; more informative under imbalance [15]), Brier score and operating-point sensitivity, specificity, positive predictive value and proportion flagged. 95% confidence intervals came from 1,000 bootstrap resamples, stratified by outcome. Paired AUC differences between the tuned logistic regression, pre-specified as the reference, and every other model were tested with De-Long’s method, [16] with Holm correction across the 13 comparisons. As a chance-level control, the tuned logistic-regression configuration was refitted 200 times on randomly permuted training labels and evaluated on the test set.

### 2.7 Data-Limitation Analyses

To ask whether the limitation lay in the data rather than the algorithm, we fitted a learning curve for logistic regression (training subsamples of 20–100%, 5 *×* 5 repeated stratified CV within the training set). We also compared three nested feature sets with tuned logistic regression and gradient boosting (Supplementary Table S5). The first was the 18 sociode-mographic candidates. The second added enrollment-interview items: age, marital status, asthma, number of pregnancies, tobacco history and preconception-counseling topics. The third added information available at delivery discharge: cesarean or operative vaginal delivery, diabetes, treatment for depression or anxiety before or during pregnancy, preterm birth and small for gestational age. Each variable’s timing was checked against the codebook. Two hypertensive-disorder items (CMAE01, A09A03b) are recorded on the same chart abstraction as the outcome without a time window, so they may capture postpartum onset; they were excluded from the discharge set and examined only in a sensitivity analysis. These models used all variables with missing-value indicators, without RFE.

### 2.8 Post Hoc Search Over Variables Recorded Before Discharge

After the main results were known, we asked whether any information recorded in nuMoM2b before delivery discharge carried more signal than the pre-specified feature sets. Candidates were the codebook variables from the screening, enrollment, antenatal-visit, ultrasound and laboratory forms, together with the sections of the delivery chart abstraction covering admission, delivery, medications and laboratory results before discharge. Sections whose time windows could extend past discharge were excluded, as were items whose labels referred to dates, discharge, the postpartum period or readmission. Inside each training fit, variables with more than 50% missing values or a minority category below 0.5% were removed, as was any variable whose value or missingness alone predicted the outcome with a univariate ROC-AUC of 0.70 or more, as a guard against leakage. An L1-penalized logistic regression, with its penalty chosen by 5-fold cross-validation, was fitted to the remaining variables, and the stability of variable selection was estimated from 100 refits on random halves of the training set. The test set was scored once, and the model was compared with the tuned 18-variable logistic regression by DeLong’s test and with 100 refits on permuted labels. This analysis was not pre-specified, and its design changed after test results had been seen: a first run was stopped after an antenatal-only variable set had been scored on the test set, and models with recursive feature elimination and the univariate leakage guard were then added.

### 2.9 Illustrative Economic Scenarios

We asked a narrower question than whether the model is cost-effective: at what cost of an outreach contact, if any, does acting on model-predicted risk outperform the two strategies that require no model, contacting every patient or contacting no one? Contacting a patient who would otherwise be readmitted yields an expected saving *B* = *e × R*, where *R* is the cost of a readmission and *e* the proportion of reached readmissions that the contact prevents. With a contact cost *c*, the rational policy contacts a patient when her predicted risk is at least *c/B*, and the value of any strategy per patient is *B ·* TP*/n − c ·* (TP + FP)*/n*. This is decision curve analysis expressed in monetary units. [14] Contacting everyone and contacting no one have equal value when *c* equals prevalence *× B*. A model can add value only when its predicted risks separate patients on either side of *c/B*, so this is the cost at which any model has the most room to help.

We applied this rule to Platt-recalibrated test-set risks (Section 2.5), so that *c/B* is a genuine risk threshold. Inputs and their sources are listed in Supplementary Table S2. The base case used *R* = $8,100, the mean hospital cost of a postpartum readmission in the Nationwide Readmissions Database [17] inflated to 2024 dollars. We used the mean rather than the median because the expected value of a strategy depends on total cost, and readmission costs are right-skewed: the median cost of a readmission to the delivery hospital was $3,981 against a mean of $5,764 (2016 dollars). [17] Readmissions related to a hypertensive disorder of pregnancy, the condition behind the effectiveness evidence below, were not cheaper: in the same database for 2013–2014, the mean cost per woman of such readmissions within 42 days was $6,100–6,600 across hypertensive disorders (about $8,100– 8,800 in 2024 dollars), and readmission costs were similar with and without a hypertensive disorder. [2] A post-discharge contact may nevertheless prevent mainly short readmissions, so as a lower bound we repeated the base case with *R* = $5,200, the same-hospital median inflated to 2024. It used *e* = 20%, close to the reductions in readmission reported for remote blood-pressure monitoring after hypertensive disorders of pregnancy (adjusted risk ratio 0.78) [18] and for transitional-care interventions in general (pooled risk ratio 0.82). [19] Effectiveness was varied over 10–50%. The upper value is optimistic: it is supported only by evidence rated low quality, [20] and a meta-analysis of randomized trials found no significant reduction. [21] Contact costs were varied from $5 to $400, a range that includes Medicaid payments for a home visit or a physician telephone call (national means $44 and $47 per claim line). [22] For each cost we report the model’s gain over the better of the two nomodel strategies, per 1,000 deliveries, with a 95% interval from 1,000 bootstrap resamples of the test set. The model’s running cost (annualized development, EHR integration and maintenance; $2,300–4,100 per 1,000 deliveries, Supplementary Table S2) is not included in that gain; we report it separately as the amount a gain would have to exceed. These scenarios are illustrative. They inherit every limitation of the underlying model and are not a cost-effectiveness analysis of a deployable tool.

### 2.10 Post Hoc Transport to State Birth Populations

This analysis was designed after the main results were known and is exploratory. The test set reflects eight academic centers, whereas the population in which a tool would be used differs from state to state in exactly the characteristics the model relies on. nuMoM2b records no state or site, so we reweighted the test participants to resemble each state’s population of first births. Targets were the 2022–2024 distributions of singleton first births by state of residence from CDC WONDER natality data: [23] race and ethnicity (five groups), Medicaid versus other payer at delivery, education (six groups), pre-pregnancy body mass index category (five groups) and maternal age (four groups). These characteristics are recorded on both the nuMoM2b forms and the US birth certificate. Smoking was not used because the two sources define it differently. Weights were obtained by raking (iterative proportional fitting) and refitted within each of 300 bootstrap resamples of the test set. We report the weighted ROC-AUC of the tuned logistic regression’s predictions, unchanged, and its Platt-recalibrated risks were used for the economics. States whose effective number of readmissions (Kish) fell below 11 were not reported. The transport assumes that the risk of readmission for a woman with given characteristics is the same in every state as in the nuMoM2b centers, so it captures differences in case mix but not in care.

For the economics, contact for women with government insurance was priced at the state’s own Medicaid payment per claim line in 2022–2023 (T-MSIS Medicaid provider spending data; [22] national figure where a state had fewer than 1,000 lines), for an established-patient home visit (CPT 99347–99350) or a physician telephone call (99441–99443). Readmission cost was as in the base case, and effectiveness was 10%, 20% or 50%. Because commercial insurers pay more than Medicaid for both the contact and the readmission, women with other insurance were analyzed under three scenarios: Medicaid prices; commercial fee-for-service prices, with readmission cost multiplied by 2.55 and contact cost by 2.85 (commercial-to-Medicare price ratios of 2.55 for inpatient and 1.88 for professional services, [24] divided by a Medicaid-to-Medicare primary-care fee ratio of 0.66 [25]); and a salaried outreach program whose contact cost does not scale with commercial prices, with only readmission cost multiplied by 2.55. In each payer group the comparator was the better of contacting all or no women in that group. A model was judged to add demonstrable value in a state when the lower bound of the 95% interval of its gain exceeded zero; we report separately the cases in which the model flagged at least 95% of women, which amounts to contacting everyone.

### 2.11 Software and Reproducibility

All analyses were run from a single script (src/revision pipeline.py) in Python 3.11.13 with scikit-learn 1.6.1, imbalanced-learn 0.14.2, FLAML 2.7.0, AutoGluon 1.6.3 and statsmodels 0.15.0; the complete environment is listed in requirements-revision.txt. Every number in this paper is read from the files that script writes (results/revision/), and per-patient predictions are stored so that every metric can be recomputed. The economic scenarios, figures and tables are produced by separate scripts that read only those files. The code is publicly available on GitHub (release v1.0.1), [26] and an interactive explorer of the economic analyses, including the state-level analysis, is publicly available and archived on Zenodo. [27]

## 3 Results

### 3.1 Cohort

Of 8,774 participants with the readmission item recorded, 154 (1.8%) were readmitted within 14 days of delivery (Table 1). The training set comprised 6,141 participants (108 readmissions) and the test set 2,633 (46 readmissions), with no participant in both. Crude readmission rates were higher after cesarean delivery (3.4%), among participants with a recorded hypertensive disorder (3.8%) and among those who smoked 20 or more cigarettes a day before pregnancy (5.3%, 12 readmissions), and lower among college graduates (1.3%).

**Table 1.**
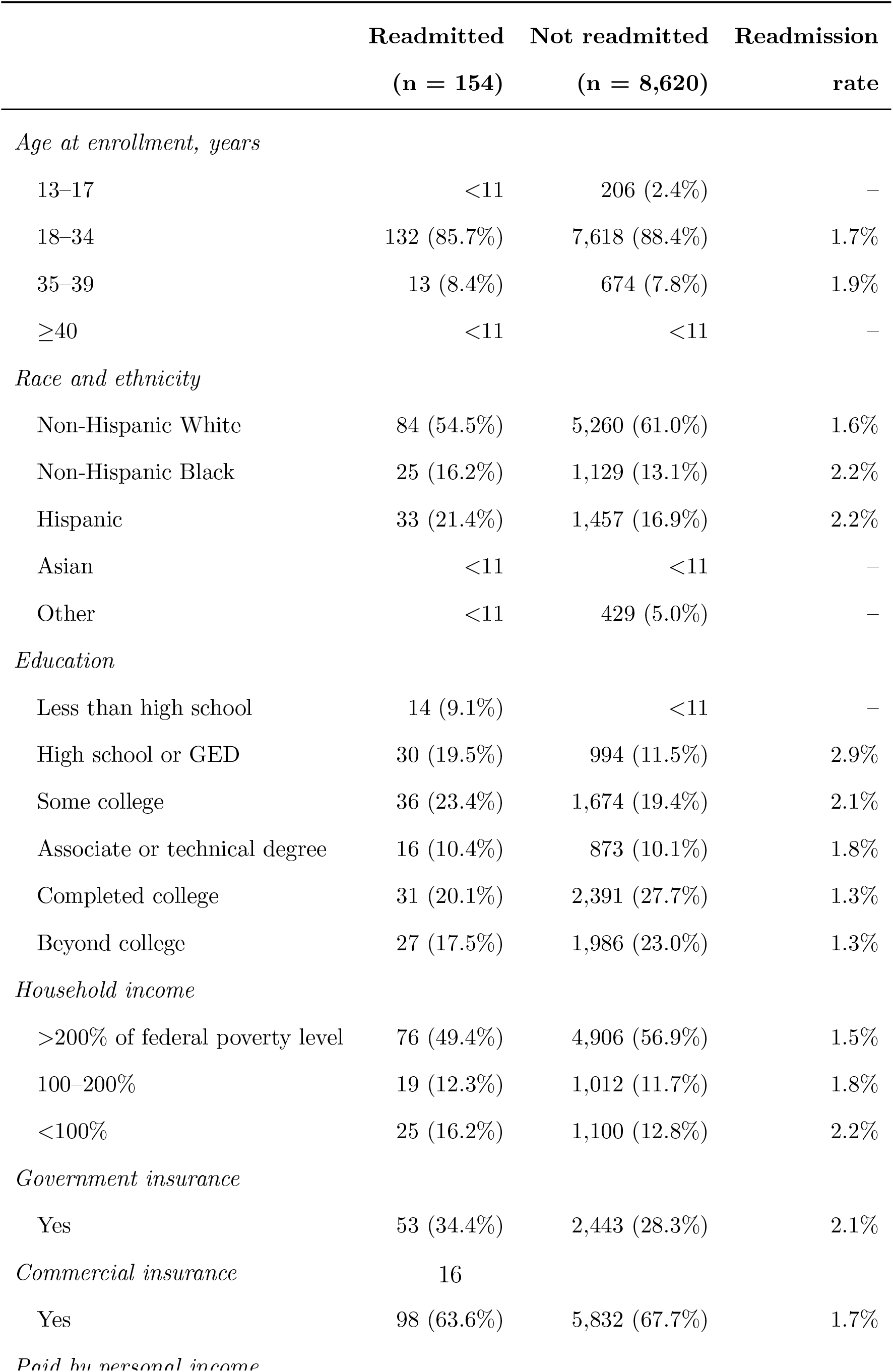
Characteristics of the analyzed cohort by 14-day postpartum readmission. Percentages in the first two columns are of each column total; the last column is the readmission rate within the category. Counts from 1 to 10 are suppressed (*<*11), with a complementary cell where needed; the two heaviest smoking categories are combined for the same reason.

|  | Readmitted | Not readmitted | Readmission |
|---|---|---|---|
|  | (n = 154) | (n = 8,620) | rate |
| *Age at enrollment, years* |  |  |  |
| 13–17 | <11 | 206 (2.4%) | – |
| 18–34 | 132 (85.7%) | 7,618 (88.4%) | 1.7% |
| 35–39 | 13 (8.4%) | 674 (7.8%) | 1.9% |
| ≥40 | <11 | <11 | – |
| *Race and ethnicity* |  |  |  |
| Non-Hispanic White | 84 (54.5%) | 5,260 (61.0%) | 1.6% |
| Non-Hispanic Black | 25 (16.2%) | 1,129 (13.1%) | 2.2% |
| Hispanic | 33 (21.4%) | 1,457 (16.9%) | 2.2% |
| Asian | <11 | <11 | – |
| Other | <11 | 429 (5.0%) | – |
| *Education* |  |  |  |
| Less than high school | 14 (9.1%) | <11 | – |
| High school or GED | 30 (19.5%) | 994 (11.5%) | 2.9% |
| Some college | 36 (23.4%) | 1,674 (19.4%) | 2.1% |
| Associate or technical degree | 16 (10.4%) | 873 (10.1%) | 1.8% |
| Completed college | 31 (20.1%) | 2,391 (27.7%) | 1.3% |
| Beyond college | 27 (17.5%) | 1,986 (23.0%) | 1.3% |
| *Household income* |  |  |  |
| >200% of federal poverty level | 76 (49.4%) | 4,906 (56.9%) | 1.5% |
| 100–200% | 19 (12.3%) | 1,012 (11.7%) | 1.8% |
| <100% | 25 (16.2%) | 1,100 (12.8%) | 2.2% |
| *Government insurance* |  |  |  |
| Yes | 53 (34.4%) | 2,443 (28.3%) | 2.1% |
| *Commercial insurance* |  |  |  |
| Yes | 98 (63.6%) | 5,832 (67.7%) | 1.7% |
| *Paid by personal income* |  |  |  |

### 3.2 Discrimination

Discrimination was modest for every model (Table 2, Figure 1). The tuned logistic regression had the highest test ROC-AUC, 0.614 (95% CI 0.542–0.685), and exceeded all 200 label-permuted refits but one (permutation *p* = 0.010; permuted AUCs 0.381–0.609). Conventional and ensemble models ranged from 0.476 to 0.614. PR-AUC did not exceed 0.026 for any model, against a test prevalence of 0.017. Hyperparameter search selected neither SMOTE nor class weighting for any of the three conventional models.

**Figure 1.**
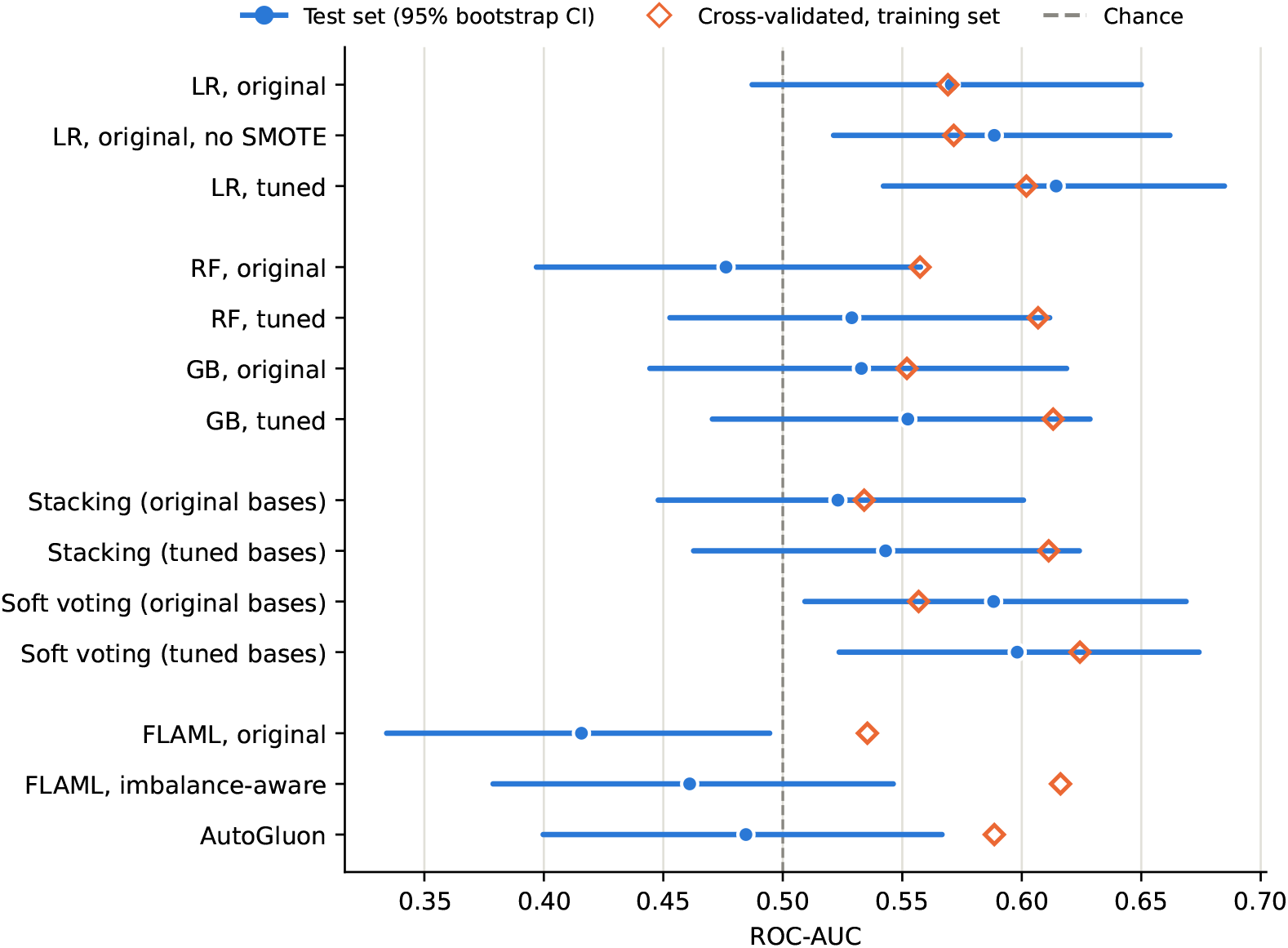
Test-set ROC-AUC with 95% stratified bootstrap confidence intervals (circles) and cross-validated training-set ROC-AUC (diamonds) for every model. Flexible models, and both AutoML frameworks in particular, lost discrimination between training and test; the tuned logistic regression did not.

**Table 2.** Performance of all models on the test set (n = 2,633; 46 readmissions). CV AUC: out-of-fold ROC-AUC on the training set (for AutoGluon, from refits within outer folds). Brier: raw, and after Platt recalibration fitted on training out-of-fold predictions; a constant prediction of the base rate scores 0.0172. Sensitivity at 0.5 is shown with the percentage of test patients flagged. Imbalance-aware FLAML is time-limited and reached a test AUC of 0.519 in a full rerun (Results). *^a^*Every model flags the same 28.4% of test patients with the highest predicted risk (the share flagged by the original logistic regression at 0.5); shown as readmissions reached of 46 and sensitivity (95% bootstrap CI).

| Model | CV AUC | Test ROC-AUC |  | PR-AUC | Brier |  | Sens. at 0.5 | Sens. at equal |
|---|---|---|---|---|---|---|---|---|
|  | (train) | Est. | 95% CI |  | Raw | Recal. | (% flagged) | workload<sup>*a*</sup> (95% CI) |
| LR, original | 0.569 | 0.571 | 0.487–0.650 | 0.022 | 0.2267 | 0.0172 | 37.0 (28.4) | 17/46, 37.0 (23.9–50.1) |
| LR, original, no SMOTE | 0.572 | 0.588 | 0.521–0.662 | 0.023 | 0.2262 | 0.0172 | 32.6 (27.7) | 16/46, 34.8 (21.7–50.0) |
| LR, tuned | 0.602 | 0.614 | 0.542–0.685 | 0.025 | 0.0171 | 0.0172 | 0.0 (0.0) | 20/46, 43.5 (30.4–56.5) |
| RF, original | 0.557 | 0.476 | 0.397–0.558 | 0.017 | 0.2312 | 0.0172 | 26.1 (29.5) | 11/46, 23.9 (10.9–34.8) |
| RF, tuned | 0.607 | 0.529 | 0.453–0.612 | 0.023 | 0.0176 | 0.0172 | 0.0 (0.0) | 15/46, 32.6 (19.6–47.8) |
| GB, original | 0.552 | 0.533 | 0.444–0.619 | 0.021 | 0.1905 | 0.0172 | 41.3 (35.6) | 16/46, 34.8 (21.7–47.8) |
| GB, tuned | 0.613 | 0.552 | 0.471–0.629 | 0.022 | 0.0175 | 0.0172 | 0.0 (0.0) | 16/46, 34.8 (21.7–47.8) |
| Stacking (original bases) | 0.534 | 0.523 | 0.448–0.601 | 0.019 | 0.2356 | 0.0172 | 41.3 (36.3) | 16/46, 34.8 (21.7–47.8) |
| Stacking (tuned bases) | 0.611 | 0.543 | 0.463–0.624 | 0.023 | 0.2439 | 0.0172 | 21.7 (20.1) | 13/46, 28.3 (15.2–41.3) |
| Soft voting (original bases) | 0.557 | 0.588 | 0.509–0.669 | 0.026 | 0.0172 | 0.0172 | 0.0 (0.0) | 18/46, 39.1 (26.0–52.2) |
| Soft voting (tuned bases) | 0.624 | 0.598 | 0.524–0.674 | 0.025 | 0.0172 | 0.0172 | 0.0 (0.0) | 19/46, 41.3 (28.3–54.4) |
| FLAML, original | 0.535 | 0.416 | 0.334–0.494 | 0.016 | 0.0359 | 0.0172 | 2.2 (1.9) | 9/46, 19.6 (8.7–32.6) |
| FLAML, imbalance-aware | 0.616 | 0.461 | 0.379–0.546 | 0.024 | 0.2139 | 0.0172 | 10.9 (10.3) | 10/46, 21.7 (10.9–34.8) |
| AutoGluon | 0.589 | 0.485 | 0.400–0.567 | 0.021 | 0.0173 | 0.0172 | 0.0 (0.0) | 12/46, 26.1 (13.0–39.1) |

Both AutoML frameworks discriminated worse than the tuned logistic regression: FLAML as originally configured 0.416 (0.334–0.494), FLAML imbalance-aware 0.461 (0.379– 0.546) and AutoGluon 0.485 (0.400–0.567). These were the only differences that remained significant after Holm correction (DeLong *p*_Holm_ = 0.003, 0.001 and 0.001); all other comparisons had *p*_Holm_ *≥* 0.075. In a full rerun of the pipeline, every other model reproduced its test AUC exactly and AutoGluon to within 0.002 (0.483, *p*_Holm_ *<* 0.001), but imbalance-aware FLAML reached 0.519 (0.444–0.598), and its difference from the tuned logistic regression was then not significant (*p*_Holm_ = 0.21). Flexible models lost discrimination between training cross-validation and the test set, whereas the tuned logistic regression did not (0.602 in CV, 0.614 on test). The losses were 0.616 to 0.461 for imbalance-aware FLAML, 0.607 to 0.529 for the tuned random forest, 0.613 to 0.552 for tuned gradient boosting and 0.611 to 0.543 for stacking of the tuned models. AutoGluon’s internal validation estimate (0.705) exceeded both its honest outer-fold estimate (0.589) and its test performance (0.485). FLAML as originally configured validated its search on a hold-out drawn from the SMOTE-resampled training data and reported an internal ROC-AUC of 0.999; the imbalance-aware configuration, validated by cross-validation on the original data, reported 0.694.

### 3.3 Default Thresholds Versus Equal Workload

At the default 0.5 threshold, sensitivity ranged from 0% to 41.3% (Table 2, Figure 2), and the spread followed how each model was trained. Models whose final estimator was trained with SMOTE or class weighting flagged 10.3% to 36.3% of test patients. The exception was FLAML as originally configured, which flagged 1.9% and reached one readmission. The tuned conventional models, the soft-voting ensembles and AutoGluon produced probabilities near the base rate and flagged no one. Sensitivity at 0.5 therefore reflects the location of a model’s probability scale, not its ability to rank patients.

**Figure 2.**
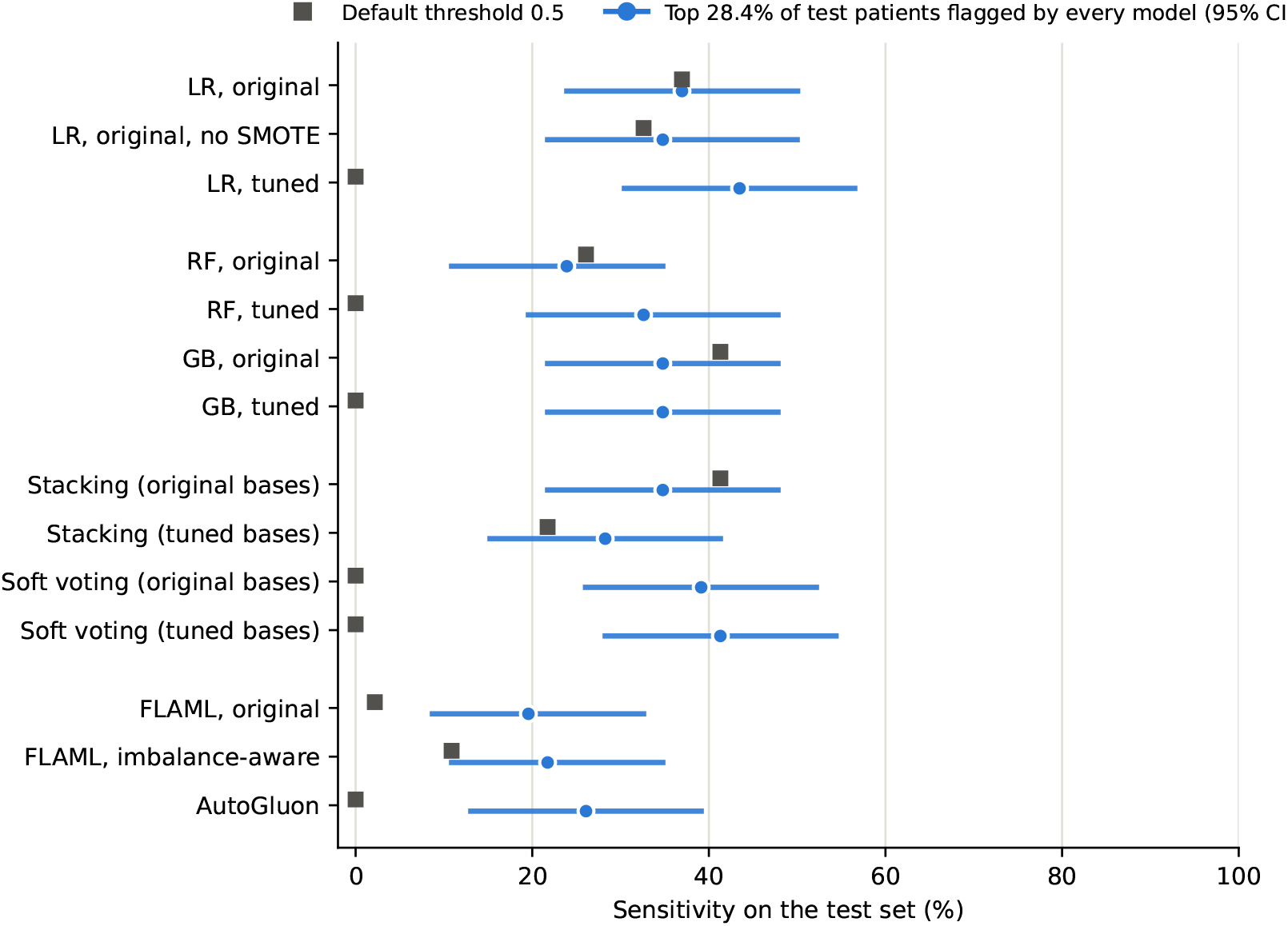
Test-set sensitivity at the default 0.5 threshold (squares) and when every model flags the same top 28.4% of test patients (circles, 95% bootstrap CI). At 0.5, sensitivity mainly reflects whether a model was trained with SMOTE or class weighting; at equal workload, conventional and ensemble models overlap, and the AutoML frameworks reach fewer readmissions.

When every model flagged the same 28.4% of test patients, sensitivity was much more uniform. Conventional and ensemble models reached 11 to 20 of 46 readmissions (23.9–43.5%), with widely overlapping intervals, and the three AutoML configurations reached 9 to 12 (19.6–26.1%). Positive predictive value was 1.2–2.7% at this workload. Thresholds chosen to flag the same proportion of training patients did not equalize workload on the test set: the share of test patients flagged ranged from 20.9% to 59.4%, because final models and out-of-fold models produced differently distributed scores. We therefore matched workload directly on the test score distribution, which uses no outcome information.

Thresholds selected on training data to reach 80% out-of-fold sensitivity achieved 58.7% test sensitivity for the tuned logistic regression, flagging 45.0% of patients. For the logistic regression as originally configured they achieved 76.1%, flagging 67.4%. Per 1,000 deliveries, the tuned model at this threshold would flag 450 women to reach 10 of the 17–18 who would be readmitted (number needed to screen 44).

### 3.4 Calibration and Decision Curves

The logistic regression as originally configured, trained with SMOTE and class weighting, greatly overestimated risk (Figure 3a). Its Brier score was 0.2267, against 0.0172 for a constant base-rate prediction, with a calibration slope of 0.34 and intercept of *−*4.04. The tuned logistic regression was close to calibrated (Brier 0.0171, slope 0.89). After Platt recalibration every model’s Brier score was 0.0172, and after isotonic recalibration 0.0172– 0.0174: no model predicted readmission better than assigning every patient the base rate. In decision curve analysis (Figure 3b), the largest net-benefit gain of any recalibrated model over the better of contacting all or no patients was 0.0022, or about two net true positives per 1,000 deliveries (tuned logistic regression).

**Figure 3.**
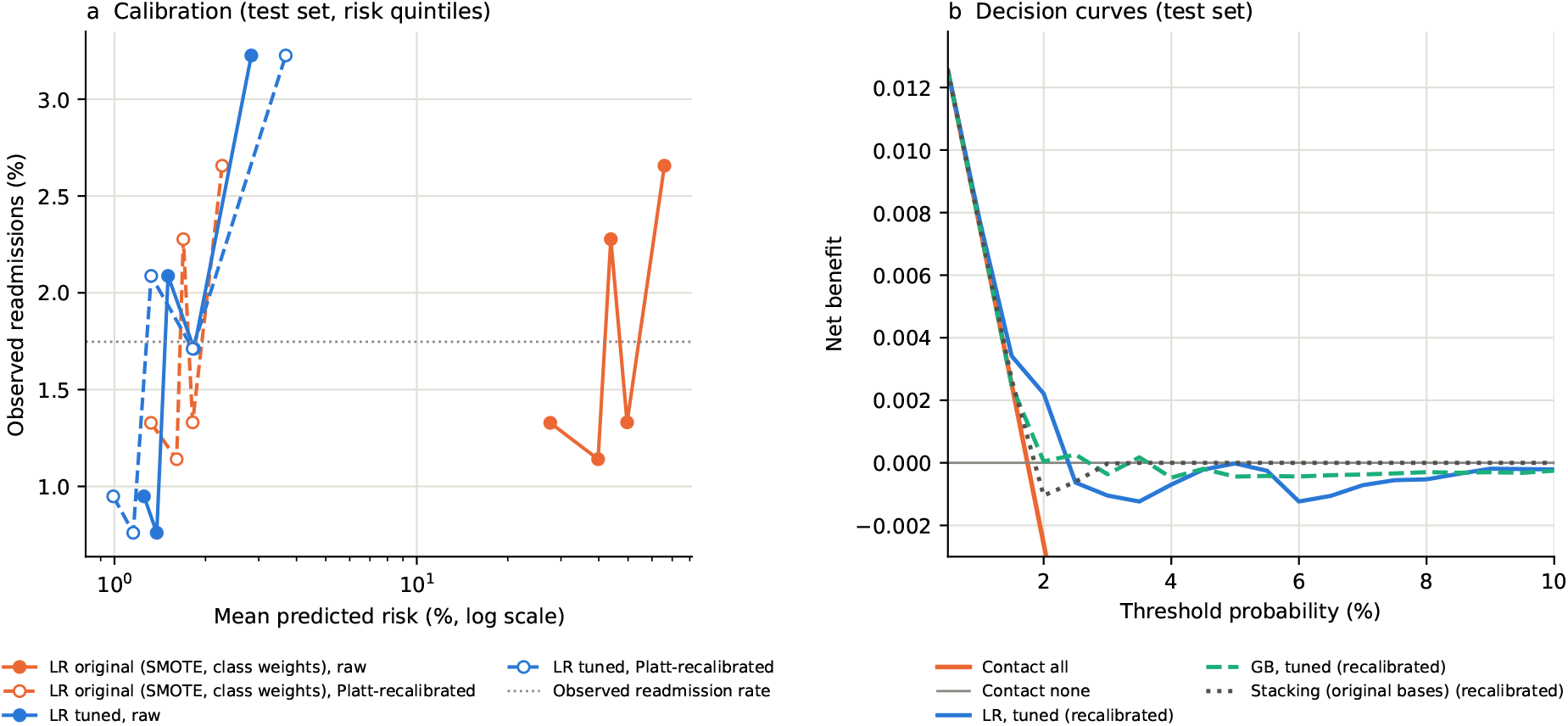
(a) Calibration on the test set by quintile of predicted risk, before (solid) and after (dashed) Platt recalibration, for the logistic regression as originally configured and as tuned. (b) Decision curves for three recalibrated models compared with contacting all or no patients.

### 3.5 Data Limitation

The pre-specified learning curve (logistic regression with RFE) stayed between 0.508 and 0.539 across training sizes (Figure 4a). This curve fell well below the same data modeled without RFE, so we added a post hoc curve for the tuned logistic regression using all 18 variables. It rose from 0.594 with 982 training participants to 0.622 with 1,964 and then remained flat (0.621–0.624), indicating that more participants with the same variables would not improve discrimination. Adding enrollment-interview items did not improve test discrimination (Table 3, Figure 4b). Adding delivery and discharge information raised test ROC-AUC to 0.611 (0.532–0.690) for logistic regression and 0.631 (0.547–0.709) for gradient boosting, with PR-AUC up to 0.034. Including the two untimed hypertensive-disorder items changed these values little (0.625 and 0.627). No feature set reached a ROC-AUC of 0.70.

**Figure 4.**
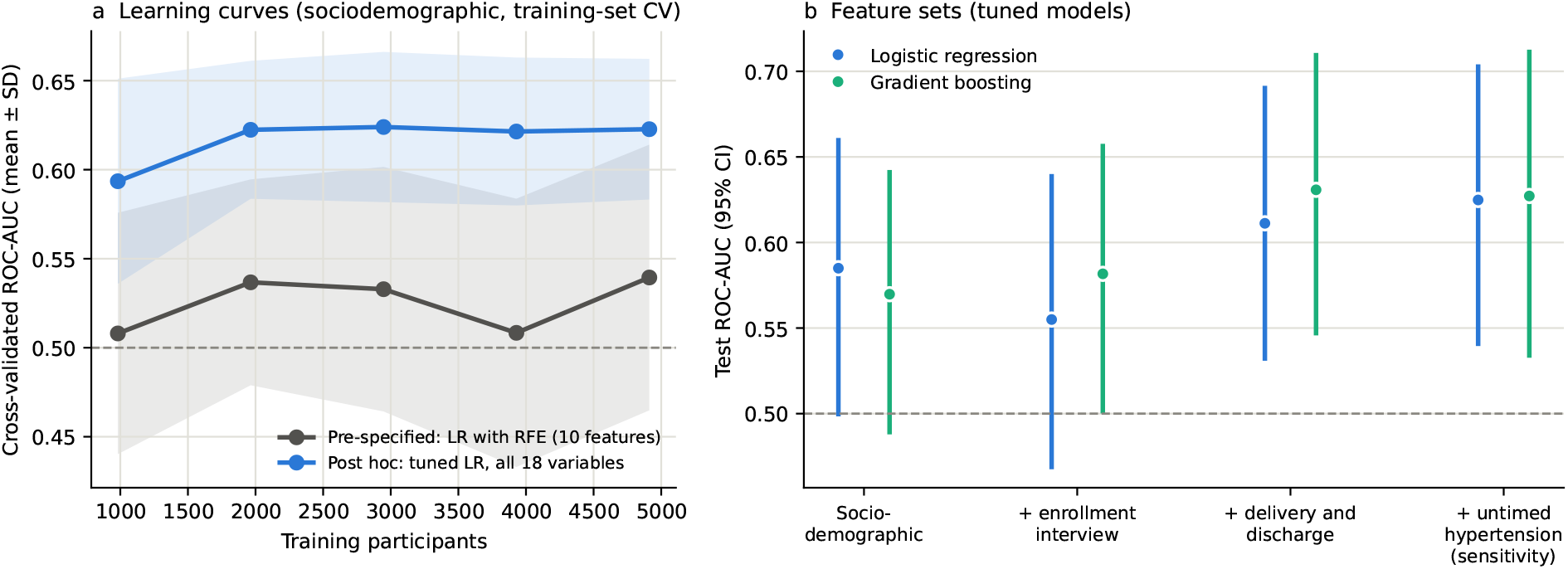
(a) Learning curves (training-set 5*×*5 repeated CV, mean *±* SD): the pre- specified logistic regression with RFE and a post hoc tuned logistic regression using all 18 sociodemographic variables. (b) Test ROC-AUC (95% CI) of tuned logistic regression and gradient boosting across nested feature sets.

**Table 3.** Discrimination by feature set (tuned models, all variables with missing-value indicators, no RFE). Variable definitions are in Supplementary Table S5.

| **Feature set** | **Model** | **Variables** | **CV AUC** | **Test ROC-AUC (95% CI)** | **Test PR-AUC (95% CI)** | **Brier** |
|---|---|---|---|---|---|---|
| Socio- demographic | Logistic regression | 18 | 0.621 | 0.585 (0.500–0.660) | 0.025 (0.019–0.040) | 0.0172 |
|  | Gradient boosting |  | 0.638 | 0.570 (0.489–0.641) | 0.023 (0.018–0.034) | 0.0173 |
| + enrollment interview | Logistic regression | 30 | 0.638 | 0.555 (0.469–0.639) | 0.022 (0.017–0.032) | 0.0174 |
|  | Gradient boosting |  | 0.643 | 0.582 (0.501–0.656) | 0.023 (0.018–0.033) | 0.0178 |
| + delivery and discharge | Logistic regression | 39 | 0.681 | 0.611 (0.532–0.690) | 0.031 (0.022–0.072) | 0.0173 |
|  | Gradient boosting |  | 0.689 | 0.631 (0.547–0.709) | 0.034 (0.023–0.057) | 0.0173 |
| + untimed hypertension (sensitivity) | Logistic regression | 41 | 0.710 | 0.625 (0.541–0.703) | 0.033 (0.022–0.079) | 0.1960 |
|  | Gradient boosting |  | 0.711 | 0.627 (0.534–0.711) | 0.029 (0.022–0.046) | 0.0175 |

In the post hoc search over variables recorded before discharge (Section 2.8), 1,370 of 4,887 candidates passed screening in the training set, and the L1-penalized model retained 110 non-zero coefficients. Its test ROC-AUC was 0.663 (95% CI 0.580–0.734; cross-validated 0.677). A full rerun of the same code reproduced these values to three decimals and selected the same twelve stable variables, but retained 113 coefficients and shifted individual selection frequencies by up to 0.03, because the L1 solver was run without a fixed random seed. This exceeded every label-permuted refit (permutation *p* = 0.010) but was not significantly higher than the tuned 18-variable model (difference 0.049, DeLong *p* = 0.24). Twelve variables were selected in at least half of the stability refits (Supplementary Table S6). They described the delivery admission and course (admission for a maternal medical or obstetric condition, cesarean delivery, other medications and prophylactic antibiotics), a history of cervical surgery, a uterine artery Doppler index, waist circumference, pre-pregnancy binge drinking, preconception counseling and tiredness in mid-pregnancy. None of the 18 sociode-mographic variables, all of which were candidates, was among the 30 most frequently selected. With 108 training events, a model with 110 coefficients is heavily over-parameterized, and only two variables were selected in more than 80% of refits, so we treat the selected variables as hypotheses rather than as a model.

### 3.6 Illustrative Economic Scenarios

At low contact costs the best strategy required no model. In the base case, a $10 contact for every patient yielded $18,302 per 1,000 deliveries, and acting on the tuned logistic regression’s recalibrated risks added $34 (95% interval $15–57) by sparing 0.3% of patients at the lowest predicted risk. The relevant comparator for a model-guided strategy is the better of contacting all or no patients, not no intervention alone.

The model’s potential gain peaked near the cost at which contacting everyone and contacting no one break even. That point is $14, $28 and $71 per contact for intervention effectiveness of 10%, 20% and 50% (Figure 5). The Medicaid payment for a home visit or telephone call ($44–47) lies above the break-even cost of the base case, so at that price contacting every patient would cost more than it saved. At $50 per contact, the tuned logistic regression contacted 10.1% of patients and its gain over contacting no one was *−*$1,341 per 1,000 deliveries (95% interval *−*$3,943 to $1,755). The largest point gains across the effectiveness range were $1,629, $3,887 and $9,692 per 1,000 deliveries, at $15, $35 and $70 respectively; none of their intervals excluded zero. At $50 no model’s interval lay above zero, and the intervals for the logistic regression as originally configured, with and without SMOTE, lay below zero (Figure 6).

**Figure 5.**
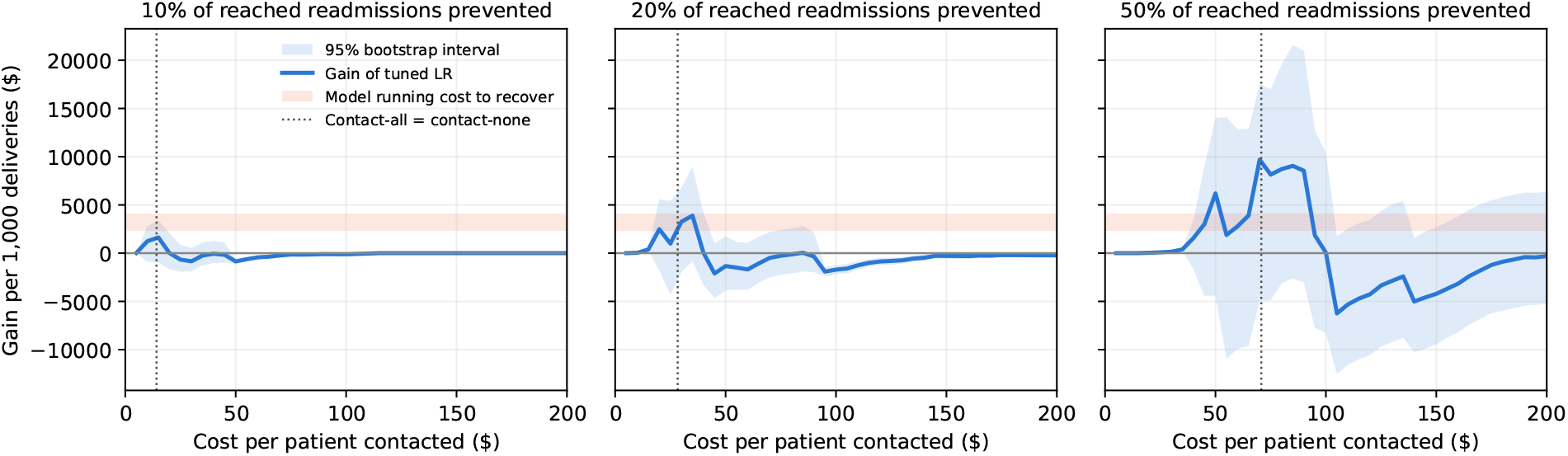
Gain of acting on the tuned logistic regression’s recalibrated risks over the better of contacting all or no patients, per 1,000 deliveries, by contact cost and intervention effectiveness (test set, n = 2,633; 46 readmissions). Blue band: 95% bootstrap interval. Dotted line: cost at which contacting all and contacting none are equally valuable. Orange band: the model’s annualized running cost, which a gain would have to exceed.

**Figure 6.**
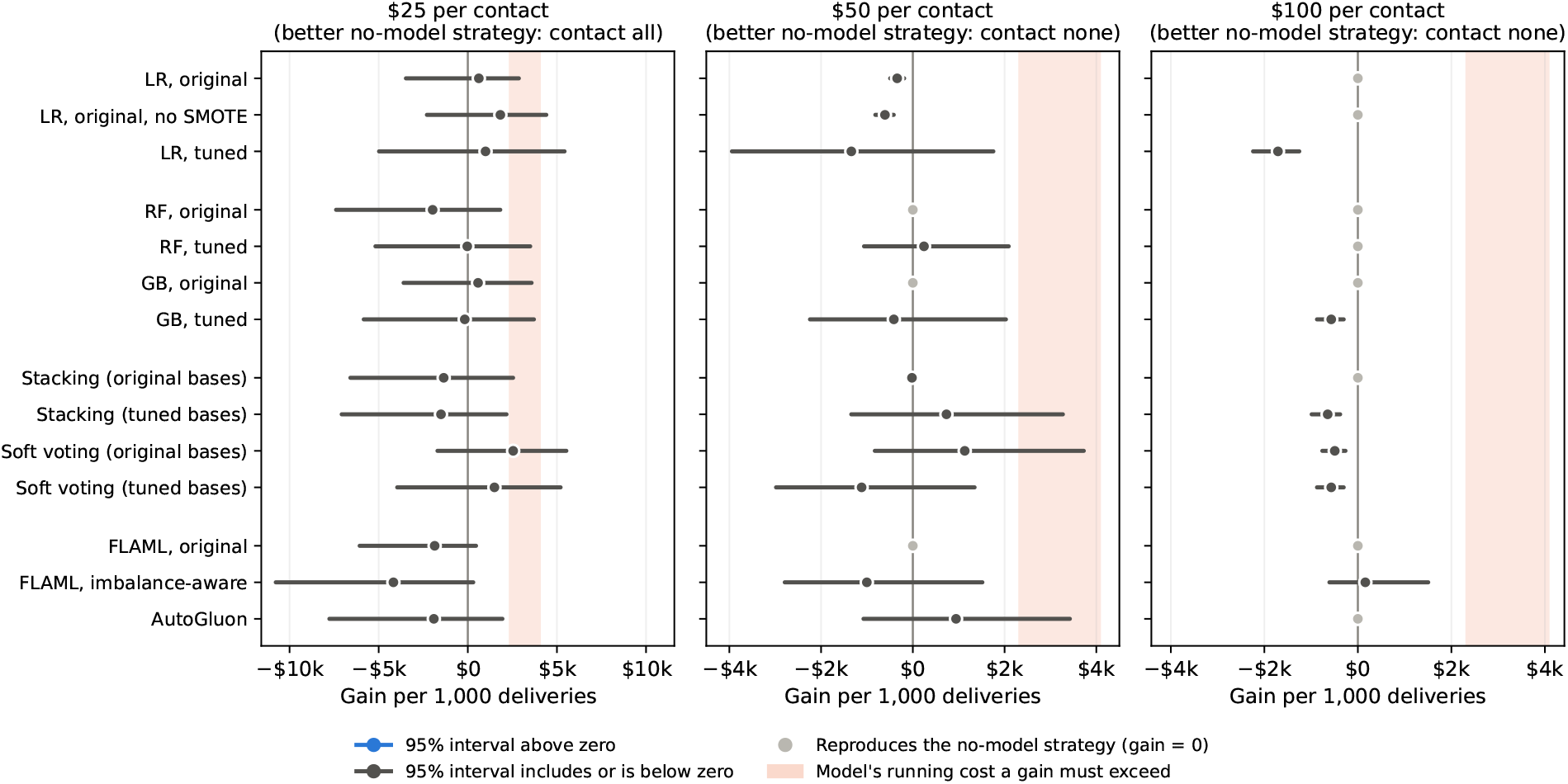
Gain of every model over the better no-model strategy at three contact costs (base case: readmission cost $8,100, 20% of reached readmissions prevented), per 1,000 deliveries, with 95% bootstrap intervals. A gain of exactly zero (gray) means that after recalibration the model gives almost every patient the same risk and therefore reproduces the no-model strategy.

Gains whose interval excluded zero appeared only just below the break-even cost, where a model withholds contact from a small group of the lowest-risk patients. For the tuned logistic regression these gains were $17 per 1,000 deliveries at 10% effectiveness, $34–382 at 20% and $38–385 at 50%. All were below the model’s running cost of $2,300–4,100 per 1,000 deliveries. Of the three models examined across the effectiveness range, the stacking ensemble, whose test discrimination did not exceed chance (Table 2), came closest. At 50% effectiveness and $50 per contact it gained $2,298 per 1,000 deliveries (95% interval $1,841– 2,659), just below the lowest running cost. Its intervals in this regime are too narrow: when the patients a model declines to contact include no observed readmission, resampling cannot generate one, so the bootstrap understates uncertainty. No model therefore showed a gain that would pay for its own running cost under any effectiveness assumption. Above the break-even cost, too few patients had predicted risks high enough to justify contact and the models lost value. With the lower readmission cost of $5,200 at 20% effectiveness, the break-even cost fell to $18 per contact, and no model’s gain had an interval excluding zero at any contact cost; the largest point gain was $2,172 per 1,000 deliveries (tuned logistic regression, $20 per contact).

### 3.7 Post Hoc Transport to State Birth Populations

Reweighting was possible for every state, and 50 jurisdictions (49 states and the District of Columbia) had enough effective readmissions to report; West Virginia was suppressed (10.2 effective readmissions). The effective sample size ranged from 448 to 2,096 of 2,633 test participants, and the largest single weight was 28. Reweighted readmission rates ranged from 1.25% to 2.74% (median 1.75%) and were higher where more first births were financed by Medicaid (correlation 0.53). Reweighted to all US first births, the rate was 1.98% and the tuned logistic regression’s ROC-AUC 0.630 (95% CI 0.544–0.712), close to its unweighted value of 0.614.

Discrimination depended on the population (Supplementary Figure S1). Transported ROC-AUC ranged from 0.531 in South Dakota to 0.667 in California (median 0.614), and its 95% interval excluded 0.5 in 30 of the 50 jurisdictions. It was lowest where first births were predominantly to non-Hispanic White women (correlation with that share *−*0.71) and highest where Hispanic (0.61) and non-Hispanic Black (0.51) women made up larger shares. The model’s predictors are sociodemographic (race and ethnicity, payer, education, income and smoking), so it separates risk only in populations in which these characteristics vary.

Model-guided contact was rarely the demonstrably better choice. At the base-case 20% effectiveness, no jurisdiction showed a demonstrable model gain at Medicaid or commercial fee-for-service prices, and one (Florida) did, for telephone calls, under the salaried-program scenario. At Medicaid prices, neither the model nor contacting everyone had a demonstrable value in 44 (home visit) and 37 (telephone) of the 50 jurisdictions, and contacting everyone was the best demonstrable strategy in 6 and 11; under the salaried-program scenario, contacting everyone was best in 18 and 23. At 10% effectiveness the model added demonstrable value nowhere. Model gains appeared mainly at the optimistic 50% effectiveness, in 0 to 4 jurisdictions per combination of contact type and price scenario, and in a further 3 to 16 the model added value only by flagging at least 95% of women. Across all 18 combinations, Georgia showed a demonstrable model gain in four, all at 50% effectiveness; no other jurisdiction did in more than one.

## 4 Discussion

### 4.1 Principal Findings

Routinely collected sociodemographic data supported only modest discrimination of 14-day postpartum readmission in this cohort of first pregnancies. The best model reached a test ROC-AUC of 0.614, and no feature set exceeded 0.631. After recalibration, no model predicted readmission better than assigning every patient the base rate, and decision curves showed no meaningful net benefit over contacting all or no patients. When models were compared at an equal screening workload, conventional and ensemble approaches were indistinguishable. Both AutoML frameworks discriminated worse, because the configurations they selected did not hold up on the test set. A model’s high sensitivity at the default threshold, set against the near-zero sensitivity of others, can look like greater clinical utility; here that contrast came from how each model’s probabilities were scaled.

### 4.2 Default Thresholds Mislead Under Class Imbalance

At the default 0.5 threshold, sensitivity ranged from zero to over 40%, and the spread was explained by whether a model had been trained with SMOTE or class weights. Both shift predicted probabilities upwards, so that 0.5 falls inside the bulk of the score distribution instead of far above it. A calibrated model of a 1.8%-prevalence outcome will rarely predict 50% for anyone, and will therefore appear to “collapse to the majority class” at that thresh-old whatever its ranking ability. The same imbalance corrections that produce apparent sensitivity also destroy calibration, as van den Goorbergh and colleagues showed for logistic regression. [11] In our data, the corrected model’s Brier score was more than ten times that of a constant prediction. Comparisons between algorithms under imbalance should therefore be made at an equal workload or across thresholds, never at the default, and probabilities intended for decisions should be recalibrated. Thresholds must also be chosen without the test set. Thresholds tuned on the test data are optimistic. Selected on training data, the threshold aimed at 80% sensitivity reached 58.7% to 76.1% on the test set, depending on the model, and required flagging 45–67% of patients.

### 4.3 Why the AutoML Frameworks Did Worse Here

With 108 training events, cross-validated estimates of discrimination are noisy, with SDs of about 0.04–0.07 in our learning curves. A search over many flexible configurations then tends to select the one whose noise happened to be favorable. AutoGluon illustrates this most clearly. Its internal estimate of 0.705 came from choosing among about 100 candidate models and fitting ensemble weights on the same out-of-fold predictions; refitting it inside outer folds gave 0.589, and the test set gave 0.485. The tuned random forest, gradient boosting and stacked ensembles showed smaller versions of the same drop, whereas the heavily regularized logistic regression did not. For FLAML as originally configured, the optimism had a second, avoidable source: oversampling the training data before the search placed synthetic readmissions interpolated from the same patients on both sides of its internal validation split, giving an internal ROC-AUC of 0.999 against 0.416 on the test set. Resampling must be applied inside each training fold only. Our conclusions concern FLAML and Auto-Gluon under the configurations and time budgets we tested, at this event count. They do not imply that AutoML is unsuitable for clinical prediction in general. They do show that, for rare outcomes, an AutoML framework’s internal validation score should not be taken at face value, and that nested or outer-fold validation is needed before its output is trusted.

### 4.4 What Data Would Be Needed

Two observations indicate that the limitation lies in the information available rather than in the algorithm or the sample size. First, once all 18 sociodemographic variables were used, the learning curve flattened from about 2,000 training participants onwards, so a larger cohort with the same variables would be unlikely to help. Second, every algorithm, tuned or not, converged on similar modest discrimination, and adding enrollment-interview items changed nothing. Information available at delivery discharge (mode of delivery, preterm birth, growth restriction and mental-health treatment) raised discrimination modestly, to 0.611–0.631. In the cohort table, readmission rates were highest after cesarean delivery and with a recorded hypertensive disorder. This points to the peripartum and early postpartum course as the most promising source of signal. A post hoc search over all variables recorded before discharge pointed the same way. Its most stable predictors described the delivery admission and course, vascular and body-size measures and mood in pregnancy, and none was sociodemographic, but even this search did not improve significantly on the 18 sociodemo-graphic variables (0.663 against 0.614). Candidate data include blood pressure trajectories and antihypertensive treatment at discharge, delivery complications and their management, length of stay, discharge laboratory values, and the social and logistical circumstances of the first postpartum week. Many of these are not captured in research cohorts designed around pregnancy outcomes. Hypertensive-disorder items whose timing could not be verified added little in our sensitivity analysis. Any future model will need the same explicit check that predictors are fixed before discharge, because postpartum hypertension is both a predictor and a cause of readmission.

### 4.5 Implications for Clinical Implementation

A screening tool built on these data would place a heavy and unproductive load on postpartum services. At an operating point aimed at capturing most readmissions, the tuned model would flag 450 of every 1,000 delivering women in order to reach about 10 of the 17–18 who would be readmitted, a number needed to screen of 44. Even at the more selective workload of 28.4%, 37 women would need to be contacted per readmission reached. Flagging close to half of a delivery population defeats the purpose of selective risk stratification, which is to concentrate scarce nursing and care-coordination time. The same arithmetic applies per flagged patient. Contacting a flagged group is worth its cost only if the proportion of that group who would otherwise be readmitted, the positive predictive value, is at least *c/B*. At the Medicaid payment for a home visit ($44) and the base-case value of $1,620 per reached readmission, that is 2.7%; at the 28.4% workload, positive predictive value was 1.2–2.7%. It also invites alert fatigue: when nearly every second discharge is flagged, staff learn to disregard the flag, including in the minority of cases where it is right. Because the strongest signals were social and economic (education, payer, smoking), flags would fall disproportion-ately on women already exposed to structural disadvantage. Without an intervention that demonstrably helps them, this risks stigmatization without benefit. Finally, probabilities from a model of a rare outcome must be recalibrated to the local population, and thresholds set and periodically reviewed by each site. That governance has a cost, and our economic scenarios suggest it would not be recovered at this level of discrimination.

#### Where a modest model could earn its keep

The economic analysis reframes the question of whether a weak model is “worth it”. The answer depends less on the model’s AUC than on where the intervention’s cost places the decision threshold relative to prevalence. When contact is cheap, contacting everyone is optimal and a screening model can only lose readmissions that it fails to flag. When contact is expensive, few patients have predicted risks high enough to justify it. Any model has room to help only near the cost at which the two no-model strategies break even, prevalence multiplied by the value of reaching one readmission. That window is narrow, and it moves with assumed intervention effectiveness (Figure 5). With a readmission cost taken from national data and effectiveness taken from the published evidence, no model in our study established a gain within this window that exceeded its own running cost. The closest, a stacking ensemble whose discrimination did not exceed chance, came near it only at the optimistic 50% effectiveness. The real prices of outreach sharpen the point: what Medicaid pays for a home visit or a telephone call lies above the base-case break-even cost, so even contacting everyone would not pay unless the intervention works better than the evidence shows. The lesson is nevertheless general.

Models with modest discrimination are routinely discarded on their AUC, yet whether a model has value depends on the intervention it would trigger. A model’s gain comes from how well it separates patients on either side of the decision threshold, which no discrimination metric captures, and the window in which this matters moves with the cost and effectiveness of the intervention. This logic applies to any rare, costly outcome. Before a risk model is built or discarded, the intended intervention’s cost and effectiveness should be used to locate this window, and candidate models should be evaluated within it. A better model would widen it.

#### Population and payer shape a model’s value

The post hoc transport to state birth populations adds a further caution. The same model, with unchanged coefficients, discriminated close to chance when the test set was reweighted to states where most first births are to non-Hispanic White women, and best when it was reweighted to more diverse states. Its signal comes from sociodemographic differences, and these vary more in some populations than in others. Its economic value moved with the case mix, the payer mix and the price of contact, and no state showed a demonstrable gain across scenarios. A model’s value is therefore a property of the model, the population and the intervention together, and an estimate from a single cohort, ours included, cannot show where deployment would pay. We present the state results as a demonstration of this dependence, not as a recommendation for or against any state. The dependence also has an equity side: a tool that discriminates mainly through race, ethnicity and payer performs best where these differences are largest, which is also where the risk of stigmatizing flags described above is greatest.

### 4.6 Comparison with Prior Work

Our findings agree with a systematic review that found no consistent performance advantage of machine learning over logistic regression for clinical prediction models, [8] and with simulation evidence that class-imbalance corrections harm the calibration of risk models. [11] They also extend a previous analysis of the same cohort, which identified demographic and socioeconomic factors associated with postpartum rehospitalization. [28] Associations of this kind can be real, as in our cohort table, without supporting useful individual prediction.

### 4.7 Limitations

#### Few events

The test set contained 46 readmissions, so every interval is wide, and single readmissions move sensitivity by more than two percentage points. We present all model variants and their intervals rather than a single best model.

#### Analyst choices made after results were seen

Five analyses were added after earlier results were available, and we label each where it is reported: the equal-workload comparison on the test score distribution, adopted after training-quantile thresholds failed to equalize workload; the learning curve without RFE; the search over variables recorded before discharge, whose design changed after test results for an antenatal variable set had been seen; the transport to state birth populations; and the two preprocessing rules (structural missingness and coding of unordered categories), adopted after earlier results had been seen. They changed individual estimates but not the conclusions.

#### Thresholds

All decision thresholds were selected on training data, but they were evaluated in a single test set and would need re-estimation at any site that adopted them.

#### Economic inputs

The economic conclusions are only as good as their inputs, and the weakest is intervention effectiveness. No trial has tested whether a post-discharge contact prevents all-cause readmission in an unselected postpartum population. The evidence we used comes from remote blood-pressure monitoring after hypertensive disorders of pregnancy, from an observational cohort and from trials that disagree, [18, 20, 21] and from transitionalcare trials outside obstetrics. [19] Our 20% base case is therefore an informed assumption, and the 50% scenario, in which the model’s window is widest, is an optimistic bound that no trial has demonstrated. The readmission cost is a mean hospital cost from 2010–2014 inflated to 2024. Hypertension-related readmissions, which remote blood-pressure monitoring targets, cost about the same as our base case, [2] but the readmissions a contact can prevent may cost less than average; a lower cost makes model-guided contact less valuable, not more, as our sensitivity analysis at $5,200 shows. What a payer pays is higher for commercially insured women, and the post hoc state analysis explores this only through national price ratios. The model’s running costs are our assumptions, and the scenarios omit indirect costs, quality of life and staff capacity. They are illustrative only.

#### State transport

Reweighting adjusts only for five characteristics recorded in both nuMoM2b and the birth certificate. It cannot capture state differences in obstetric and discharge practice, in access to care after discharge, or in the risk associated with a given profile, any of which could move a state’s result in either direction. The same characteristic can also be recorded differently on research forms and birth certificates. Women in categories that are sparse at nuMoM2b’s academic centers received large weights. The largest weights (up to 28) and smallest effective sample sizes (down to 448) occurred for Alaska and Hawaii, so results for such states rest on few participants. Rurality, which is not available in the data we analyzed, could not be adjusted for at all. Every state reuses the same 2,633 participants and 46 readmissions, so the state results are a sensitivity analysis, not independent state estimates. Commercial prices were scaled from national ratios rather than state-specific data.

#### Single cohort and outcome definition

nuMoM2b enrolled nulliparous women at academic centers, and readmission was abstracted from charts at the delivery hospital, so readmissions elsewhere may have been missed. Results may not transfer to multiparous or community-hospital populations.

#### AutoML scope

We evaluated two frameworks under specific configurations and time budgets. Other frameworks, longer searches or custom objectives might behave differently.

### 4.8 Future Directions

Prediction of postpartum readmission should be attempted with data on the delivery hospitalization and early postpartum period, collected with explicit timestamps relative to discharge, and evaluated with recalibration, equal-workload comparisons and decision analysis rather than discrimination alone. Because the evidence that post-discharge contact prevents readmission comes almost entirely from remote blood-pressure monitoring after hypertensive disorders of pregnancy, [18, 20, 21] a more promising design pairs a cause-specific outcome with an intervention known to act on it, for example hypertension-related readmission with remote blood-pressure monitoring, rather than all-cause readmission with an unstructured telephone call. The economic window described here can be used before model development, with that intervention’s cost and effectiveness, to decide whether a model is worth building at all.

## 5 Conclusions

Routinely collected sociodemographic data cannot support a useful tool for predicting 14-day postpartum readmission after a first birth, whatever the algorithm. The best model reached a ROC-AUC of 0.614 and, once recalibrated, offered no net benefit over contacting all or no patients. Apparent differences between algorithms at the default threshold reflected how class-imbalance corrections scale predicted probabilities, not differences in ranking ability. Two AutoML frameworks discriminated worse than a regularized logistic regression because their internal validation was optimistic at this event count. Delivery and discharge information added a little signal, and the learning curve was flat, so future work should prioritize richer peripartum and postpartum data over larger samples of the same variables. Whether any model is worth deploying depends on the cost and effectiveness of the intervention it would trigger, and on the population: for cheap outreach, contacting everyone is the better policy, and only within a narrow cost range can a weakly discriminating model add value. In our data no model recovered its running cost there, and discrimination varied with the population to which the model was applied. Decision-economic evaluation in the intended population should therefore precede the decision to build, deploy or discard a modest model.

## Data Availability

Complete code and analysis pipelines are available in the project repository, enabling full replication of results. 18. Frasch, Martin Wakefield, Colin (2023). ML model for prediction of postpartum rehospitalization in pregnant women/new mothers using readily obtainable pre-pregnancy or early pregnancy sociodemographic and health determinants. figshare. Software. https://doi.org/10.6084/m9.figshare.23573373.v5

## Acknowledgments

The authors thank the nuMoM2b investigators and the study participants whose data made this research possible.

## Use of generative AI

The authors used Claude (Anthropic), a generative AI assistant, to help write and review analysis code, check that every reported number traces to a result file, search for and verify literature, and edit the text. The authors reviewed the code, results and text and take full responsibility for the content.

## Author contributions (CRediT)

**Colin Wakefield:** Methodology, Formal analysis, Data curation, Writing – review & editing. **Ciprian P. Gheorghe:** Validation, Supervision, Writing – review & editing. **Martin G. Frasch:** Conceptualization, Methodology, Software, Formal analysis, Supervision, Project administration, Writing – original draft, Writing – review & editing. All authors read and approved the final manuscript.

## Ethical Statement

This study is a secondary analysis of de-identified data from the nuMoM2b study. The parent nuMoM2b study was approved by the institutional review boards of all participating clinical sites, and all participants provided written informed consent. [10] The present secondary analysis used a de-identified, access-controlled research dataset, did not involve any new contact with human participants, and accessed no protected health information; it was therefore exempt from additional institutional review board review as non-human-subjects research. The work was conducted in accordance with the Declaration of Helsinki.

## Data Availability Statement

The nuMoM2b dataset is not publicly available without restriction. It is held by the *Eunice Kennedy Shriver* National Institute of Child Health and Human Development (NICHD) Data and Specimen Hub (DASH) and is available to qualified investigators through a data use agreement and controlled-access application (https://dash.nichd.nih.gov). The authors accessed the data under such an agreement and are not permitted to redistribute participant-level data. The derived variable specifications and the code needed to reproduce the results from the data are provided in the code repository.

## Code Availability

All analysis code is available at https://github.com/martinfrasch/numom2b-postpartum-risk-prediction-health-economics (release v1.0.1). [26] It includes the candidate-variable specification, the temporal variable classification used to prevent leakage, and the single pinned modeling script (src/revision pipeline.py). The repository also contains the economic analysis (src/economic check.py), the scripts that generate every figure and table from the result files, and the frozen software environment (requirements-revision.txt). With access-approved nuMoM2b data, the repository reproduces every reported number. The repository contains no data, predictions or result files, in keeping with the data use agreement. An interactive explorer of the economic analyses is available at https://martinfrasch.github.io/postpartum-readmission-economics/ and archived on Zenodo (doi:10.5281/zenodo.23749950). [27] It uses only grouped results, each group containing at least 11 readmissions and 11 non-readmissions, so its values approximate those reported here.

## Supplementary Materials

### Supplementary Table S1: Variable Temporal Classification

**Table S1.** nuMoM2b Variable Classification by Temporal Relationship to Readmission Outcome.

| **Tier** | **Dataset Prefixes** | **Count** | **Description** |
|---|---|---|---|
| A (Safe) | a02–a35, demographics, v1a–v1l | 1,068 | Pre-pregnancy, enrollment, Visit 1 |
| B (Safe) | v2a–v4a, v2b–v3l, vxx | 2,115 | Prenatal visits 2–4 |
| C (Borderline) | cma*, cmb, cmc, cmd, cba–cbc, cla–clb | 3,116 | Delivery and peripartum |
| D (Leakage) | cme, CMAJ* | 132 | Post-discharge complications and readmission |
| Other | sleep, food, physical activity, etc. | 5,186 | Non-clinical research data |
| **Total** |  | **11,617** |  |

\*CMA dataset contains both delivery variables (Tiers A–C) and readmission variables (CMAJ prefix = Tier D). CMAE-prefix variables within CMA require individual review: some capture pre-pregnancy history (safe), others capture postpartum conditions (leakage).

### Supplementary Table S2: Health-Economic Assumptions

**Table S2.** Health-economic assumptions used in the illustrative scenarios (inputs, not results), with their sources. All costs in 2024 US dollars.

| Parameter | Base case | Range | Source and derivation |
|---|---|---|---|
| *Costs* |  |  |  |
| Cost of a postpartum readmission (*R*) | \$8,100 | \$5,200–11,100 | Nationwide Readmissions Database 2010–2014, all payers, readmissions within 60 days: mean hospital cost (charges converted with cost-to-charge ratios) $5,764 when readmitted to the delivery hospital and $8,499 elsewhere, in 2016 dollars. [17] Base case: mean weighted by the number of readmissions to each, inflated by the consumer price index (CPI-U, 2016 to 2024, factor 1.307). Mean, not median, because expected value depends on total cost. Upper bound: the different-hospital mean, inflated. Lower bound: the same-hospital median ($3,981), inflated, as a proxy for the shorter readmissions a contact can plausibly prevent. Hypertension-related readmissions within 42 days in the same database, 2013–2014: mean $6,100–6,600 per readmitted woman across hypertensive disorders (cost-to-charge converted; about \$8,100–8,800 in 2024 dollars, CPI-U factor 1.336), similar to readmissions without a hypertensive disorder. [2] A hospital-cost perspective; payer payments differ (below). |
| Cost per contact (*c*) | varied | \$5–400 | Swept. Reference points, Medicaid paid amount per claim line, 2022–2023, national (T-MSIS): [22] home visit, established patient (CPT 99347–99350) $44; physician telephone call (99441–99443) $47; transitional care management (99495, 99496) $88 and $90. State values vary widely (home visit, state median \$40). |
| Commercial-to-Medicare price ratio | 2.55; 1.88 | – | Inpatient facility and professional services, 2022 claims. [24] Used only in the post hoc state analysis. |
| Medicaid-to-Medicare fee ratio | 0.66 | – | Primary care, 2024 national index. [25] Used only in the post hoc state analysis. |
| Model development and validation | \$35,000 | \$25,000–50,000 | Authors' assumption (one-time); we found no published per-model estimate. |
| EHR integration and training | \$20,000 | \$15,000–30,000 | Authors' assumption (one-time). |
| Model maintenance | \$20,000/yr | \$15,000–25,000 | Authors' assumption (monitoring, recalibration). |
| Running cost per 1,000 deliveries | \$3,000 | \$2,300–4,100 | Development and integration amortized over 5 years plus maintenance, at 10,000 deliveries per year. |
| *Clinical* |  |  |  |
| Intervention effectiveness (*e*), relative reduction in readmission among patients contacted | 20% | 10–50% | No trial has tested post-discharge contact against all-cause readmission in an unselected postpartum population. Remote blood-pressure monitoring after hypertensive disorders of pregnancy: adjusted risk ratio 0.78 (0.65–0.93) for 6-week readmission in a propensity-matched cohort of 12,038; [18] odds ratio 0.4 (0.23–0.71) in a meta-analysis of trials rated low quality; [20] no significant reduction in hypertension-related readmission in a meta-analysis of four trials (5.3% vs 11.8%). [21] Transitional-care trials outside obstetrics: pooled risk ratio 0.82 (0.73–0.91) for 30-day readmission. [19] 50% is an optimistic upper bound. |
| Readmission rate | 1.8% | – | nuMoM2b analyzed cohort (14 days); state-transported rates in Section 3.7. |
| *Population* |  |  |  |
| US births, 2024 | 3,628,934 | – | Final natality data; 1,427,020 (39.3%) were first births. [29] |

### Supplementary Table S3: Variables Excluded as Outcome Leakage

**Table S3.** Variables excluded because they encode or follow the readmission outcome.

| **Variable** | **Form** | **Reason for Exclusion** |
|---|---|---|
| CMAJ01d2 | CMA | Readmission reason (preeclampsia) — directly encodes outcome |
| CMAJ01A_INT | CMA | Days to readmission — outcome timing variable |
| CMAJ01b_hrs | CMA | Hours hospitalized on readmission — outcome variable |
| CMAJ01c | CMA | Number of readmissions — outcome variable |
| CMAJ01d1–d5 | CMA | Readmission reasons — outcome components |
| CMAE10 | CMA | Severe anemia requiring transfusion — postpartum complication |
| CMAE11, CMAE11a1–b3 | CMA | Blood product transfusion details — postpartum |
| CMAE04a1c | CMA | Depression – postpartum (concurrent with outcome) |
| CMAE04a2c | CMA | Anxiety – postpartum (concurrent with outcome) |

*Note: Pre-pregnancy versions of CMAE variables (e.g., CMAE04a1a “Depression – Prior to pregnancy”) are temporally safe but were collected on the same form, requiring careful extraction.*

### Supplementary Table S4: Hyperparameter Search Spaces and Selected Values

**Table S4.**
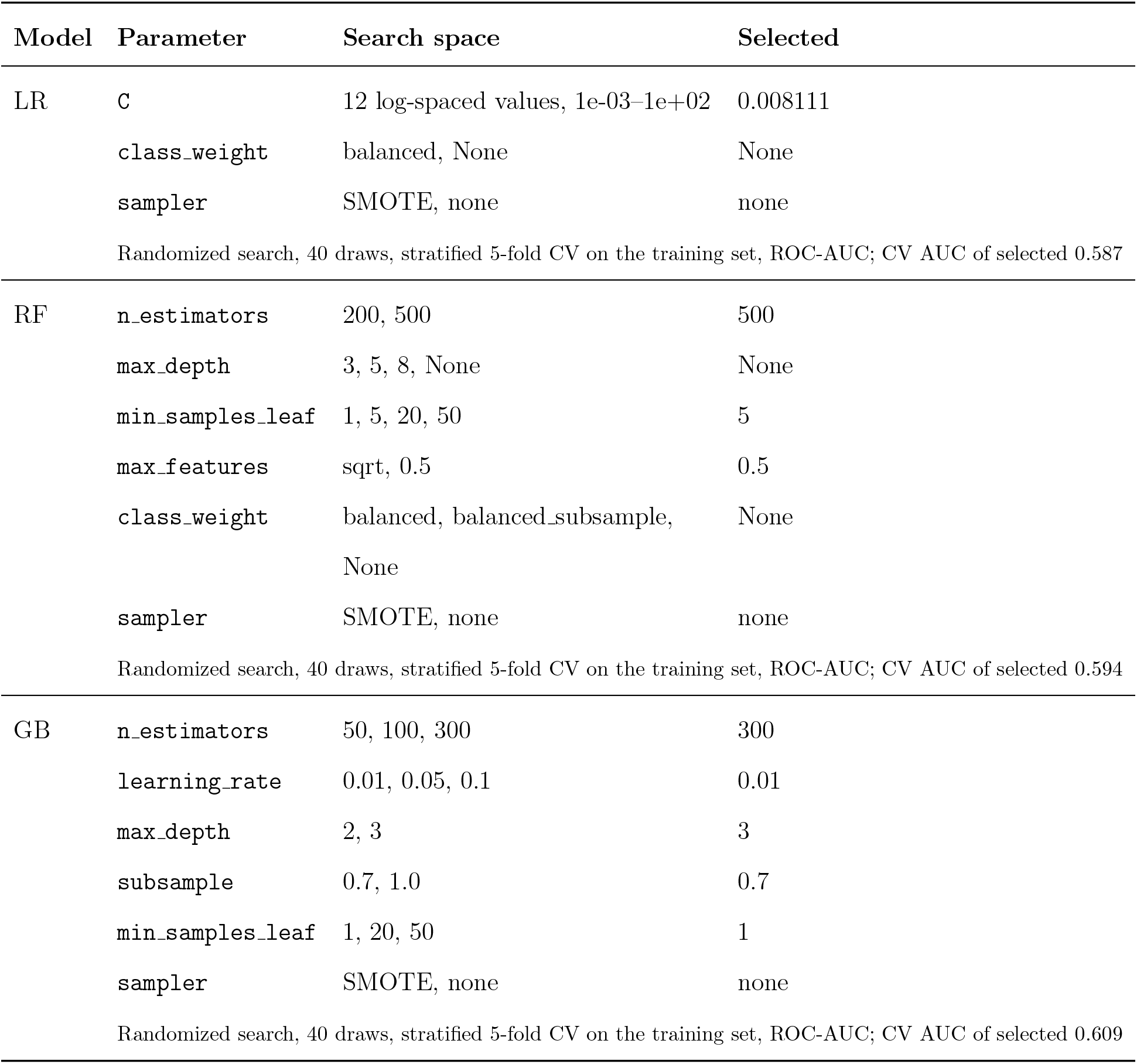
Hyperparameter search spaces and the values selected for the tuned models. sampler: SMOTE or none, tuned jointly with class weighting.

| Model | Parameter | Search space | Selected |
|---|---|---|---|
| LR | C | 12 log-spaced values, 1e-03–1e+02 | 0.008111 |
|  | class_weight | balanced, None | None |
|  | sampler | SMOTE, none | none |
|  | Randomized search, 40 draws, stratified 5-fold CV on the training set, ROC-AUC; CV AUC of selected 0.587 |  |  |
| RF | n_estimators | 200, 500 | 500 |
|  | max_depth | 3, 5, 8, None | None |
|  | min_samples_leaf | 1, 5, 20, 50 | 5 |
|  | max_features | sqrt, 0.5 | 0.5 |
|  | class_weight | balanced, balanced_subsample, None | None |
|  | sampler | SMOTE, none | none |
|  | Randomized search, 40 draws, stratified 5-fold CV on the training set, ROC-AUC; CV AUC of selected 0.594 |  |  |
| GB | n_estimators | 50, 100, 300 | 300 |
|  | learning_rate | 0.01, 0.05, 0.1 | 0.01 |
|  | max_depth | 2, 3 | 3 |
|  | subsample | 0.7, 1.0 | 0.7 |
|  | min_samples_leaf | 1, 20, 50 | 1 |
|  | sampler | SMOTE, none | none |
|  | Randomized search, 40 draws, stratified 5-fold CV on the training set, ROC-AUC; CV AUC of selected 0.609 |  |  |

### Supplementary Table S5: Feature Sets for the Data-Limitation Analysis

**Table S5.** Feature sets used in the data-limitation analysis. Each set includes all variables of the sets above it. Timing relative to discharge was verified against the nuMoM2b codebook.

| **Set** | **Variables (nuMoM2b name: description)** |
|---|---|
| Sociodemographic (primary) (18) | CRace: race/ethnicity (5 groups); Race: race/ethnicity (8 groups); eRace: race (NIH categories); eHispanic: Hispanic ethnicity; BMI: BMI at enrollment; BMI_Cat: BMI category; Education: education; GravCat: gravidity category; SmokeCat1: ever used tobacco; SmokeCat2: smoked in 3 months before pregnancy; SmokeCat3: cigarettes per day (none if not smoking); Ins_Govt: government insurance; Ins_Mil: military insurance; Ins_Comm: commercial insurance; Ins_Pers: paid by personal income; Ins_Othr: other payer; PctFedPoverty: income, % of federal poverty level; poverty: poverty category |
| Added: enrollment interview (12) | Age_at_V1: age; AgeCat_V1: age category; V1AF04: marital status; V1AD15: asthma; V1AE01: number of pregnancies; V1AG04: ever used tobacco (interview); V1AG05: smoked in 3 months before pregnancy (interview); V1AD02a: counseled: folic acid; V1AD02b: counseled: healthy weight; V1AD02c: counseled: vaccines; V1AD02d: counseled: dental visit; V1AD02e: counseled: genetic counseling |
| Added: delivery and discharge (9) | CMAC15: cesarean delivery; CMAC12: operative vaginal delivery attempted; CMAE03: diabetes ever diagnosed; CMAE04a1a: treated for depression before pregnancy; CMAE04a1b: treated for depression during pregnancy; CMAE04a2a: treated for anxiety before pregnancy; CMAE04a2b: treated for anxiety during pregnancy; A09A03a: preterm birth (<37 weeks); SGA_alex: small for gestational age |
| Added in sensitivity analysis only (timing not recorded) (2) | CMAE01: hypertension, proteinuria or related condition; A09A03b: suspected maternal hypertensive disorder |

### Supplementary Table S6: Stability Selection in the Post Hoc Search

**Table S6.** Post hoc, exploratory. Variables recorded before delivery discharge that the L1-penalized logistic regression selected in at least half of 100 refits on random halves of the training set. Selection frequency is not an effect size, and the direction of association depends on each item’s coding.

| **Variable** | **Description** | **Frequency** |
|---|---|---|
| V3EA02b | Visit 3 (22–29 wk): bothered by tiredness or low energy | 0.89 |
| CMAA01m | Admitted to labor and delivery for a maternal medical or obstetric condition | 0.85 |
| CMAC15 | Cesarean delivery | 0.68 |
| V1AD12c | Previous cervical surgery: cryotherapy | 0.68 |
| U2CD05 | Right uterine artery Doppler, systolic/diastolic ratio | 0.67 |
| V1AD02a | Preconception counseling: folic acid | 0.67 |
| V1AD12b | Previous cervical surgery: LEEP | 0.64 |
| V1AD12a | Previous cervical surgery: cone biopsy | 0.61 |
| V1AG02c | Episodes of four or more drinks, three months before pregnancy | 0.60 |
| V1BA04b | Waist circumference (visit 1) | 0.59 |
| CMAG06i | Other prescribed medication during the delivery stay | 0.55 |
| CMAG02i1 | Other prophylactic antibiotic during the delivery stay | 0.53 |

### Supplementary Figure S1: Discrimination Transported to State Birth Populations

**Figure S1.**
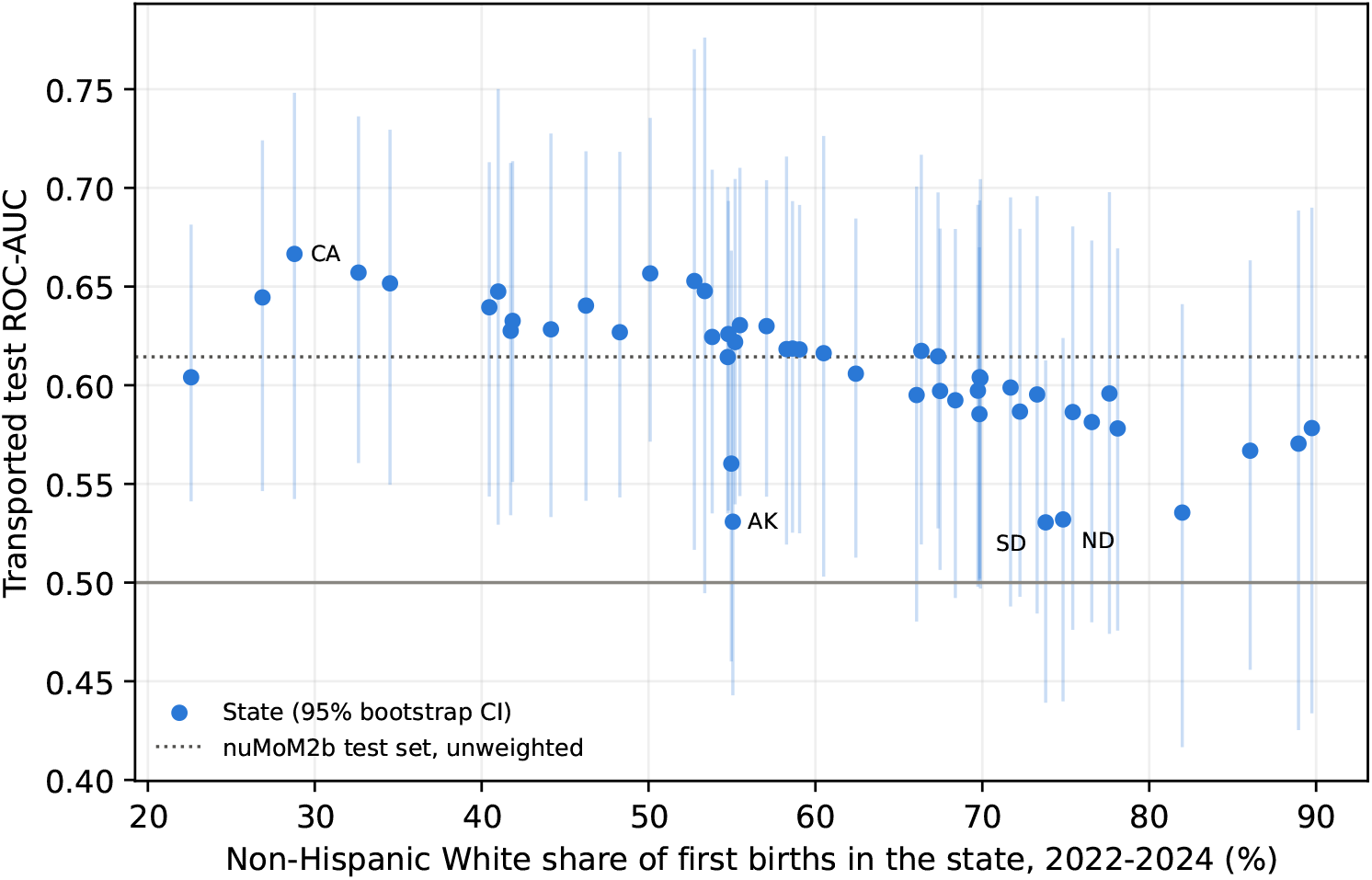
Post hoc, exploratory. ROC-AUC of the tuned logistic regression on the test set reweighted to each jurisdiction’s 2022–2024 singleton first births (CDC WONDER), against that jurisdiction’s non-Hispanic White share of first births. Bars: 95% bootstrap intervals with weights refitted in each resample. Dotted line: unweighted test ROC-AUC. Labeled: the lowest three and the highest. West Virginia is not shown (*<*11 effective readmissions).

